# Redefining typhoid diagnosis: what would an improved test need to look like?

**DOI:** 10.1101/19001958

**Authors:** Richard Mather, Heidi Hopkins, Christopher M. Parry, Sabine Dittrich

## Abstract

**Introduction:** Typhoid fever is one of the most common bacterial causes of acute febrile illness in the developing world, with an estimated 10.9 million new cases and 116.8 thousand deaths in 2017. Typhoid point-of-care (POC) diagnostic tests are widely used but have poor sensitivity and specificity, resulting in antibiotic overuse that has led to the emergence and spread of multidrug resistant strains. With recent advances in typhoid surveillance and detection, this is the ideal time to produce a target product profile (TPP) that guides product development and ensure that a next-generation test meets the needs of users in the resource-limited settings where typhoid is endemic.

**Methods:** A structured literature review was conducted to develop a draft TPP for a next-generation typhoid diagnostic test with minimal and optimal desired characteristics for 36 test parameters. The TPP was refined using feedback collected from a Delphi survey of key stakeholders in clinical medicine, microbiology, diagnostics and public and global health.

**Results:** A next-generation typhoid diagnostic test should improve patient management through the diagnosis and treatment of infection with acute Salmonella enterica serovars Typhi or Paratyphi with a sensitivity ≥90% and specificity ≥95%. The test would ideally be used at the lowest level of the healthcare system in settings without a reliable power or water supply and provide results in less than 15 minutes at a cost of <$1.00 USD.

**Conclusion:** This report outlines the first comprehensive TPP for typhoid fever and is intended to guide the development of a next-generation typhoid diagnostic test. An accurate POC test will reduce the morbidity and mortality of typhoid fever through rapid diagnosis and treatment and will have the greatest impact in reducing antimicrobial resistance if it is combined with diagnostics for other causes of acute febrile illness in a treatment algorithm.

## INTRODUCTION

Typhoid fever (typhoid) is an enteric bacterial infection caused by *Salmonella enterica* serovar Typhi (*Salmonella* Typhi; *S*. Typhi). It is one of the most common bacterial causes of acute febrile illness in the developing world,[1] with an estimated 10.9 million new cases worldwide and 116.8 thousand deaths in 2017.[2,3] Paratyphoid fever caused by *Salmonella enterica* serovars Paratyphi A, B and C (*S*. Paratyphi) results in a disease that can have an identical clinical syndrome to typhoid fever,[4] but is often less severe.[5] Typhoid fever is most common in South Asia and sub-Saharan Africa, with children predominantly affected. Like many febrile illnesses, typhoid presents with non-specific symptoms and signs, especially in its early stages. In routine healthcare settings in low- and middle-income countries (LMIC), typhoid fever is commonly suspected and treated empirically with antibiotics.[6] This overuse of antibiotics creates a selective pressure for the development of antimicrobial resistance (AMR),[7] that has resulted in the emergence and spread of typhoid strains that are resistant to all first-line antibiotics.[8] Similarly, the low specificity of current rapid diagnostic tests (RDTs) can lead to an over diagnosis of typhoid fever that may result in the overuse of antibiotics and delay the proper treatment for underlying conditions. For example, the use of the Widal test during an outbreak of acute febrile illness in Nepal led to misdiagnosis of typhoid which delayed the appropriate treatment of the causative agent (scrub typhus), resulting in dozens of deaths.[9] The potential harms of current typhoid RDTs are compounded by the fact they are widely available, cheap and easy to use.[10]

Various aspects of *S*. Typhi biology make diagnosis by standard laboratory methods challenging. *S*. Typhi is able to bypass the gastrointestinal mucosal barrier that restricts other enteric bacteria and can evade the typical innate immune responses with limited activation of inflammatory pathways.[11] *S*. Typhi infection begins with invasion of the mucosa of the terminal ileum, and the organism is thought to only be transiently present in the blood before dissemination throughout the reticuloendothelial system into the bone marrow, liver and spleen.[12] The bacterial load in peripheral blood peaks in the first week of illness,[13] but is still very low with a median of 0.1-1.0 colony forming units (CFU)/mL in symptomatic patients.[14] This concentration is difficult to detect by blood culture or PCR, resulting in lower sensitivity for these diagnostic tests. *S*. Typhi is a member of the Enterobacteriaceae family, and antibodies that have been produced in response to prior infections with other Enterobacteriaceae tend to cross-react with *S*. Typhi,[12] due to significant conservation of surface antigens. This cross-reactivity lowers the specificity of antibody-based diagnostic assays that otherwise are well suited for a simple, rapid and inexpensive test format. Compounding the challenge, the muted immune response that occurs through expression of the Vi capsular polysaccharide[12] may further hinder the utility of serological tests for typhoid diagnosis.

In clinical settings supported by a microbiology laboratory, invasive typhoid infection is confirmed through isolation of *S*. Typhi from blood cultures, but this is relatively expensive, can take >48 hours, has low sensitivity, and requires laboratory infrastructure and trained staff that are not commonly available in LMIC where typhoid is most prevalent.[15] Bone marrow cultures have high sensitivity for detection of *S*. Typhi but are not routinely used because of the invasive techniques needed to obtain bone marrow aspirates. PCR testing for typhoid is expensive and has a low diagnostic sensitivity when used on peripheral blood samples.[16] Other available point-of-care (POC) diagnostic tests include the Widal test,[17] TUBEX,[18] Typhidot,[19] Test-it Typhoid,[20] and the Typhoid-Paratyphoid diagnostic assay (TPTest). However, these tests all have significant drawbacks that limit their clinical use.

A Cochrane review of the accuracy of the commercially available antibody-based rapid RDTs showed moderate sensitivity and specificity for the TUBEX colorimetric test that detects anti-O:9 antibody titres (78%, 87%), the Typhidot dot enzyme linked immunosorbent assay (ELISA) that measures IgG and IgM antibodies against the outer membrane proteins of *S*. Typhi (84%, 79%) and the Test-it Typhoid immunochromatographic lateral flow assay that detects IgM antibodies against *S*. Typhi O antigen (69%, 90%).[21] The TPTest is a newer serological test that detects circulating IgA using ELISA with a sensitivity and specificity of >95%.[22] But it takes 24-48 hrs hours to produce a result, and requires blood culture equipment not widely available in resource-limited settings.[14,22] Due to the limited sensitivity of all current typhoid POC tests they cannot be relied upon to guide treatment prescribing.

If developed and implemented effectively, an accurate typhoid RDT could reduce morbidity and mortality through faster diagnosis. Further, it could help to reduce the overuse of antibiotics that contributes to the emergence and spread of multidrug resistant strains of *S*. Typhi and other bacteria. In recent years, novel approaches have been described to develop typhoid diagnostic tests with improved accuracy in resource-limited settings, including serological, molecular, metabolomic, proteomic, and transcriptomic methods.[7] For example, a recent study has shown that IgA and IgM against *S*. Typhi lipopolysaccharide (LPS) may be a specific marker of acute typhoid infection and is a promising target for diagnostic test development.[23]

As the need for appropriate fever case management becomes more apparent,[24] the need for improved typhoid diagnostics suitable for use in low-resource environments becomes more pressing. Building on the recent momentum around improved typhoid surveillance[25] and advances in typhoid detection,[1] this report describes the development of a target product profile (TPP) in an attempt to define the diagnostic needs for this important pathogen. The TPP is intended to guide product development and to ensure an optimized solution that meets the needs of endemic countries and results in tangible improvements in patient management. In addition, the TPP aims to re-invigorate the discussion of diagnostics as a crucial part of the global typhoid agenda. This report focuses on the process of TPP development with an emphasis on key test characteristics and discussion points identified by typhoid experts and experienced stakeholders.

## METHODS

### Data gathering

A structured review of relevant literature related to *Salmonella* Typhi diagnosis was performed to develop a draft TPP with minimal and optimal desired characteristics for a next-generation typhoid diagnostic test. The test characteristics chosen for the TPP were selected based on previous TPPs published by the Foundation for Innovative New Diagnostics (FIND)[26,27], and include the scope, target population, intended use, expected test performance, as well as operational and financial parameters (Table 1). Each desired test characteristic was classified as either a minimum requirement that a test must meet to be useful for healthcare providers treating patients in resource-limited settings, or an optimum threshold that would make the test highly desirable for both healthcare providers and patients. The Ovid Medline database was accessed on June 21, 2018 using the Medical Subject Heading (MeSH) “Typhoid fever” and the subheading “Diagnosis”. Results were restricted to English language articles published in the previous 10 years. Titles and abstracts of retrieved articles were scanned for relevance, with articles of interest thoroughly reviewed by RM for content relevant to the TPP. Additional documents were identified by searching for “typhoid” on the websites of the Cochrane library, WHO, and FIND, and by screening references and studies that cited articles selected in the initial search. Expert stakeholders to be contacted for the Delphi survey were identified as part of the literature review.

**Table 1:** Typhoid target product profile parameters

| Category | Test characteristic |
| --- | --- |
| Scope of test | Goal<br>Target population<br>Target user<br>Target level of health system |
| Test performance | Sample type<br>Sample collection<br>Sample volume/ sample transfer device<br>Additional sample preparation<br>Ease of use<br>Hands on time<br>Time to result<br>Read out of results<br>In use stability<br>Data output + connectivity<br>Data interpretation<br>Analyte type<br>Multiplexing<br>Analytical sensitivity/ Limit of detection (LoD)<br>Diagnostic sensitivity<br>Diagnostic specificity<br>Reproducibility |
| Operational characteristics | Kit configuration<br>Reagent preparation<br>Operating conditions<br>Transportation and storage stability<br>Equipment (Instrumentation external to test)<br>External maintenance<br>Calibration<br>Internal/ Process control<br>Batch/Quality control<br>Power requirements<br>Water requirement<br>Waste disposal<br>Bio-safety<br>Training requirements<br>Cost per test |

### Delphi survey

Stakeholders were contacted for input on the draft TPP using a Delphi survey. Stakeholders included specialists in clinical medicine (n = 14), laboratory medicine (n = 2), microbiology (n = 6), diagnostics (n = 11) and public health and global health. An online survey (supplementary table 1) was used and respondents were asked to rate their agreement with each of the TPP characteristics using a Likert scale (1 = strongly disagree, 2 = disagree, 3 = neither agree nor disagree, 4 = mostly agree, 5 = fully agree). A consensus agreement was defined as ≥75% of respondents who either mostly or fully agreed with a TPP characteristic. Results from the first round of the survey (Oct/2018) were used to refine the TPP, and a second draft of the TPP was distributed (Nov/2018) to all initial participants as well as two additional stakeholders identified after the first round of the survey was completed.

## RESULTS

An Ovid Medline search using the MeSH “Typhoid fever” produced 10,698 results, with 1,558 results for the subheading “Diagnosis”. Limiting search results to English language articles published after January 1^st^, 2008, provided 298 articles that were screened for relevance. Additional documents were included as outlined in the Methods and selected articles were thoroughly reviewed to develop a draft typhoid TPP with minimum and optimum criteria for the test characteristics in Table 1.

Feedback on the draft TPP was obtained from key stakeholders through the first round of the Delphi survey, with 40 stakeholders contacted and 19 (19/40, 48%) completed surveys received. Survey respondents had experience working in low resource settings in Africa, the Americas, Europe, the Eastern Mediterranean, South-East Asia and the Western Pacific region. Consensus agreement of ≥75% was achieved for 34/36 (94%) TPP criteria. TPP criteria that generated the most discussion in the Delphi survey were related to the scope of the test including the goal, target population, level of the health care system, diagnostic sensitivity and specificity, as well as cost. Based on feedback from survey respondents, “multiplexing” was removed as a TPP characteristic, and the remaining minimum and optimum TPP criteria were revised. For criteria that had achieved consensus agreement, revisions were made if survey respondents provided compelling suggestions for improvement.

A second draft of the TPP was distributed to the 19 people who responded to the initial Delphi survey, and two additional stakeholders identified after completion of the first round. A total of 12 completed surveys were received from 13 stakeholders, including two who submitted a joint survey, with consensus agreement achieved for 33/35 (94%) TPP characteristics. The two criteria that did not meet the consensus threshold were the target level of the health system and diagnostic sensitivity, both of which received 67% agreement. Survey respondent feedback was used to revise these two criteria, and to make minor changes to four criteria that had ≥75% agreement, before inclusion in the final version of the TPP presented in Tables 2, 3 and 4.

**Table 2.** Typhoid target product profile characteristics: scope of the test

| Characteristic | Minimal requirement | Optimal requirement | References |
| --- | --- | --- | --- |
| Goal | Point-of-care test to improve patient management through diagnosis and treatment of infection with acute *Salmonella enterica* serovars Typhi or Paratyphi | Combine with diagnostics for malaria and other causes of acute febrile illness as part of a treatment algorithm | [8,30] |
| Target population | All individuals with undifferentiated acute fever |  | [15,25,28] |
| Target user | Laboratory technician | Healthcare worker | [7] |
| Target level of health system* | District hospital with basic laboratory facilities | Primary health posts and centres | [6,29] |

\* Consensus not reached among survey respondents.

**Table 3:**
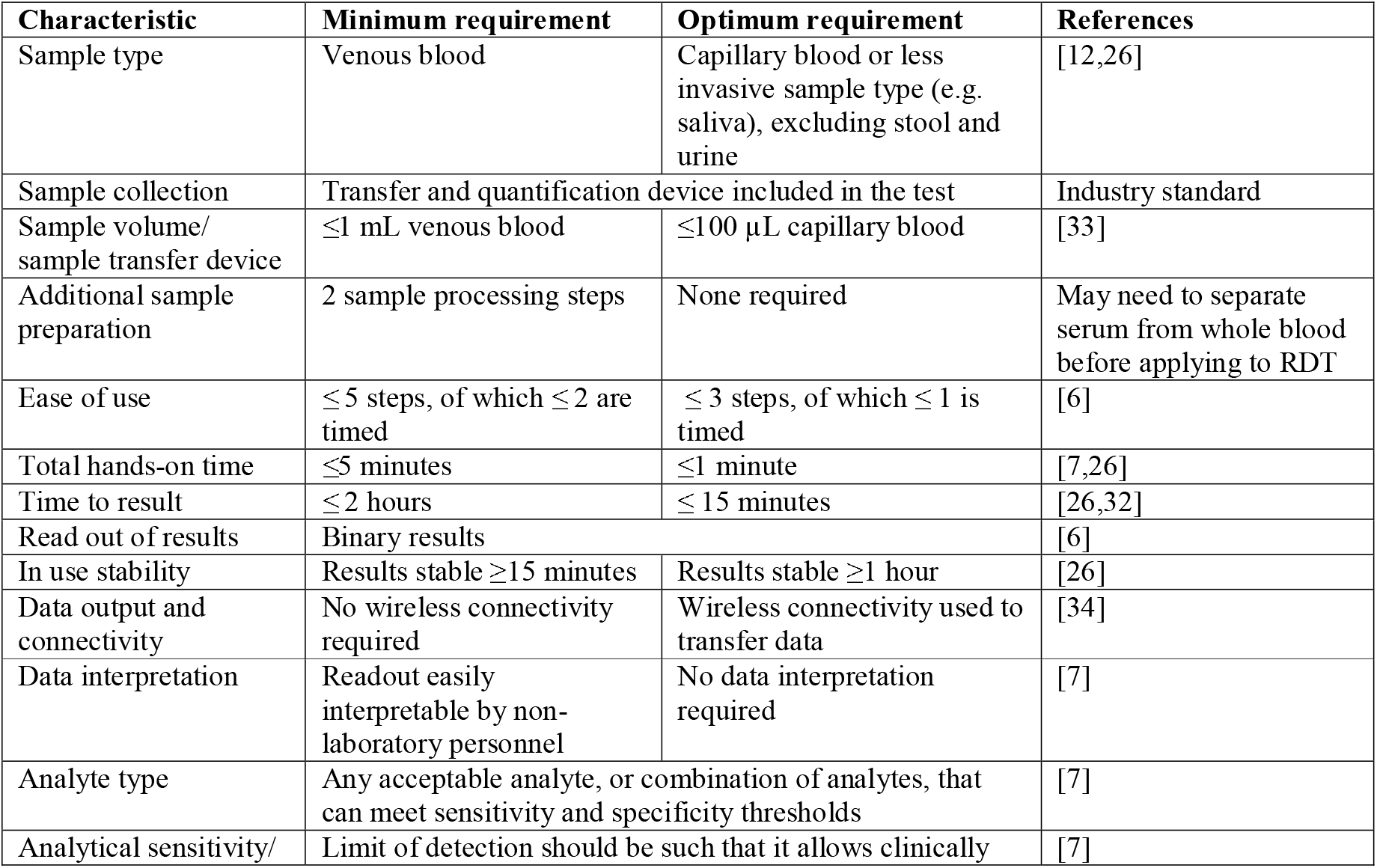
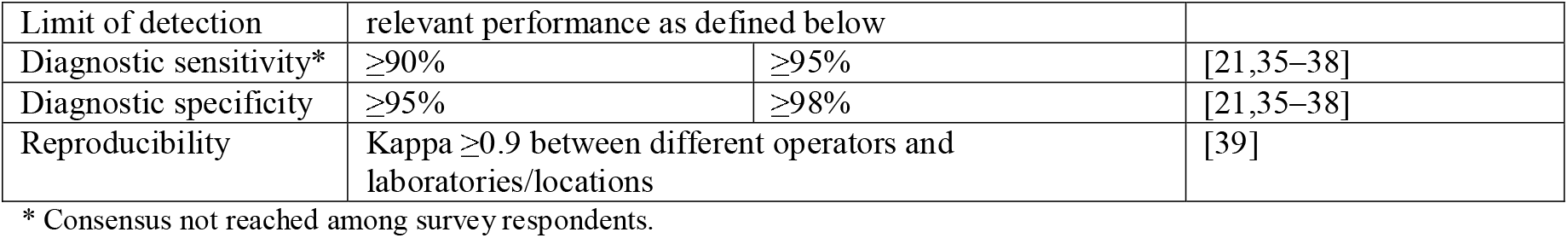
Test performance characteristics for a typhoid diagnostic target product profile

| Characteristic | Minimum requirement | Optimum requirement | References |
| --- | --- | --- | --- |
| Sample type | Venous blood | Capillary blood or less invasive sample type (e.g. saliva), excluding stool and urine | [12,26] |
| Sample collection | Transfer and quantification device included in the test |  | Industry standard |
| Sample volume/<br>sample transfer device | $\leq 1$ mL venous blood | $\leq 100$ μL capillary blood | [33] |
| Additional sample preparation | 2 sample processing steps | None required | May need to separate serum from whole blood before applying to RDT |
| Ease of use | $\leq 5$ steps, of which $\leq 2$ are timed | $\leq 3$ steps, of which $\leq 1$ is timed | [6] |
| Total hands-on time | $\leq 5$ minutes | $\leq 1$ minute | [7,26] |
| Time to result | $\leq 2$ hours | $\leq 15$ minutes | [26,32] |
| Read out of results | Binary results |  | [6] |
| In use stability | Results stable $\geq 15$ minutes | Results stable $\geq 1$ hour | [26] |
| Data output and connectivity | No wireless connectivity required | Wireless connectivity used to transfer data | [34] |
| Data interpretation | Readout easily interpretable by non-laboratory personnel | No data interpretation required | [7] |
| Analyte type | Any acceptable analyte, or combination of analytes, that can meet sensitivity and specificity thresholds |  | [7] |
| Analytical sensitivity/ | Limit of detection should be such that it allows clinically |  | [7] |
| Limit of detection | relevant performance as defined below |  |  |
| Diagnostic sensitivity* | ≥90% | ≥95% | [21,35–38] |
| Diagnostic specificity | ≥95% | ≥98% | [21,35–38] |
| Reproducibility | Kappa ≥0.9 between different operators and laboratories/locations |  | [39] |

\* Consensus not reached among survey respondents.

**Table 4:** Consensus operational characteristics for the typhoid target product profile

| Characteristic | Minimum requirement | Optimum requirement | References |
| --- | --- | --- | --- |
| Kit configuration | Package of single kits sharing reagents (if required) and user manual. Instructions in English, French, Spanish and Portuguese. | Package of single kits with individual reagents (if required) sharing user manual. Instructions in local languages. | [26,40] |
| Reagent preparation | One reagent preparation step | None required | [26,41] |
| Operating conditions | - between 5 and 40°C<br>- ≤90% relative humidity | - between 5 and 45°C<br>- ≤90% relative humidity | [40] |
| Transportation and storage stability | ≥ 12 months at ≤35°C and ≤70% relative humidity, no cold chain needed, ability to withstand transport stress (≤3 days at 60°C) | ≥ 24 months at ≤45°C and ≤90% relative humidity, no cold chain needed, ability to withstand transport stress (≤3 days at 60°C) | [40] |
| Equipment | Small, portable or handheld, battery- | No equipment | [6,7] |
| (Instrumentation external to test) | operated instrument |  |  |
| External maintenance | Minimal maintenance, simple to perform by non-laboratory personnel | No maintenance | [26] |
| Calibration | ≤1 annual calibration, ideally auto-calibration by operator or remotely | No calibration | [26] |
| Internal/ Process control | Included in each assay |  | Industry standard |
| Batch/Quality control | Positive and negative controls included in each kit |  | Industry standard |
| Power requirements | Battery or solar powered | No power needed | [26] |
| Water requirement | No external water required |  | [26] |
| Waste disposal | Biohazard waste, sharps disposal | No toxic waste requiring special disposal | [42] |
| Bio-safety | Basic biosafety level 1, WHO Class B In-vitro diagnostic (moderate individual and low public health risk) | Basic biosafety level 1, WHO Class A In-vitro diagnostic (low individual and low public health risk). | [42,43] |
| Training requirements | ≤ 0.5 days for lab technician | ≤ 0.5 days for experienced health worker | [6,7,21] |
| Cost per test | End-user cost <\$3.00 USD | End-user cost <\$1.00 USD | [7,26] |

### Scope of test

Delphi survey feedback emphasized that a next-generation RDT for typhoid fever should not focus solely on the diagnosis of *S*. Typhi. To reduce the empiric use of antibiotics that generates selective pressure for AMR, a typhoid RDT would ideally be combined with diagnostics for malaria and other causes of acute febrile illness as part of a case management algorithm (Table 2). Due to the similar clinical presentation and the changing epidemiology of *S*. Typhi and *S*. Paratyphi, survey respondents advised that a next-generation test for typhoid fever should be able to detect both *S*. Typhi and *S*. Paratyphi.

The target population was identified based on published data from Africa and Asia.[25,28] Children aged two to 14 years bear the brunt of the global typhoid burden but there is substantial variability both within and between regions in terms of who is most affected. A recent study from Pakistan found higher rates of typhoid fever in adults than in children.[28] These data highlight the need for a typhoid RDT that can detect the disease in individuals of all ages, as confirmed by the Delphi survey.

When designing new diagnostic tests, the level of the healthcare system where a test will be deployed is an important consideration. Ghani, *et al*., have identified five healthcare system levels, with different types of diagnostic or prognostic tools suitable for different levels.[29] Typhoid is most prevalent in LMIC with limited healthcare resources, and in these contexts the optimal typhoid test would not require sophisticated equipment and could be easily interpreted by non-laboratory personnel.[6,7] Respondents agreed that a test would optimally be usable at the lowest level of a healthcare system, which in many cases is a community health worker seeing patients in an informal environment. However, as the current gold standard of blood culture requires laboratory equipment, but has suboptimal sensitivity and specificity, some respondents felt it was acceptable for a typhoid RDT to require basic laboratory facilities, with a trained laboratory technician, providing it meets all other TPP criteria. Based on feedback from the initial round of the Delphi survey the minimum target level was adjusted upward to a higher level of the health care system, but consensus agreement was not achieved as some respondents felt strongly that the minimum requirement should be a test that can be used in informal settings at the lowest level of the healthcare system.

### Test performance

Blood culture is commonly used as the reference standard for typhoid diagnosis but requires sophisticated equipment not readily available in LMIC where typhoid is endemic.[6] Typhoid blood cultures require a minimum of two to 10 mL of venous blood due to the low bacterial load in peripheral blood, and have poor sensitivity estimated at only 61% in a recent systematic review.[31] The most commonly used typhoid POC tests (Widal, Typhidot, Tubex, Test-It Typhoid, TPTest) require between 5 µL and 1 mL of blood, but have only moderate sensitivity and specificity.[21] Survey respondents agreed that an optimal next-generation typhoid RDT would use a capillary blood sample with a volume of ≤100 µL, or a less invasive sample type, excluding urine and stool[12,26]. However, survey respondents indicated that ≤1 mL of venous blood was an acceptable minimum requirement due to the current difficulty in accurately diagnosing typhoid fever (Table 3). The TPP allows for up to two sample processing steps as an RDT may require serum to be separated from whole blood, with at most five steps for the test of which no more than two should be timed,[26] and a total hands on time of less than five minutes. Based on a published expert consensus TPP for diagnostics for acute febrile illness, RDT performance ideally would entail three or fewer steps, of which at most one step is timed, with a total hands-on time of one minute or less.[26]

For a new typhoid diagnostic test to have the greatest impact on prescribing and clinical outcomes it would need to yields results in less than a few hours.[32] There was consensus agreement that the optimum requirement would make results available within 15 minutes to coincide with the average development time of other point-of-care diagnostics commonly used in LMIC environments (for example, malaria antigen-detecting RDTs). A minimum requirement of results within two hours was agreed; this would be a significant improvement from the ≥48 hours required for blood culture, and two hours was deemed the longest time that outpatients could wait for test results, particularly in rural settings where patients may have to travel long distances to reach a health facility.[26]

Researchers have proposed that an ideal typhoid diagnostic test would have a simple positive/negative read-out similar to a home pregnancy test,[6] with results easily interpretable by non-laboratory personnel.[7] The typhoid TPP therefore requires a binary read-out of results, with data that either do not require interpretation, or that are easily interpretable. No specific analyte or limit of detection is specified for the typhoid TPP, with any analyte or combination of analytes acceptable providing the test meets all other TPP requirements.

The minimum TPP requirement for diagnostic sensitivity is ≥90%, with an optimum sensitivity of ≥95%, based on modelling data and expert opinion.[21,35–38] Consensus agreement in the Delphi survey was not achieved for test sensitivity, which reflects the substantial variation in published expert opinions regarding the desirable accuracy for a typhoid RDT. However, survey respondents did agree on a minimum specificity of 95% and an optimum specificity of 98%.

### Operational characteristics

Survey respondents agreed on operational characteristics of the typhoid TPP (Table 4). Typhoid diagnostic test kits ideally should consist of individually packaged tests with individual reagents (if required) and a user manual in local languages, based on TPP characteristics for other POC tests in regions where typhoid is endemic.[26,40,41] Up to one reagent preparation step is acceptable, to allow for reconstitution of a powdered reagent. The test should not require a cold chain, with operating conditions that reflect the high temperatures and humidity that are present in many regions in Africa and Asia where typhoid is prevalent.

Currently most typhoid treatments are provided in outpatient settings, including informal medical shops, so an ideal POC test would not require any sophisticated equipment or a formal laboratory infrastructure.[7] A small, portable or handheld battery-operated instrument is acceptable,[44] but ideally no equipment would be required. To be truly transformative, a typhoid POC test needs to be useable in settings without a reliable power or water supply. If power is required, then it should be provided by a combination of rechargeable batteries and solar power.

Empiric treatment of suspected typhoid cases is common, typically using relatively inexpensive antibiotics.[7] To reduce the overuse of empiric antibiotics, the end-user cost for a typhoid POC test was set at <$3.00 USD (minimum requirement) or <$1.00 USD (optimum requirement) to reflect the cost of empiric antibiotics in endemic regions.[7] Delphi survey feedback indicated that the highest cost to the end-user in Africa should be equivalent to one US dollar.

## DISCUSSION

Typhoid diagnostic tests currently lack the sensitivity and specificity required for an accurate diagnosis at the point of care, resulting in the overuse of antibiotics through empiric treatment. The WHO has developed a list of characteristics that make a test suitable for the resource-limited settings where typhoid is prevalent: the ASSURED acronym stands for affordable, sensitive, specific, user-friendly, rapid and robust, equipment-free, and delivered to those in need.[44] TPPs build upon these criteria and increasingly are used in the global health community to guide development of diagnostic tests and to inform donors about global health priorities.[26,40,41] This TPP outlines the minimum and optimum desired characteristics for an improved typhoid RDT and is intended to accelerate development of optimized diagnostics that meet the needs of users in endemic regions.

Antibiotic resistance is a growing threat to typhoid treatment, with strains of *S*. Typhi that are resistant to three first-line agents now prevalent in parts of Asia and Africa.[8] The emergence of multi-drug resistant strains that have acquired additional resistance to fluoroquinolones and third-generation cephalosporins, known as extremely drug resistant *S*. Typhi, has left azithromycin and the costly intravenous carbapenem drugs as the only antibiotic options for some patients.[8] There have been sporadic case reports of azithromycin-resistant *S*. Typhi,[8] but if extremely drug resistant strains acquire azithromycin resistance, carbapenems could be left as the only effective treatment. To prevent the further spread of resistant *S*. Typhi it would be beneficial to conduct drug susceptibility testing for individual patients before commencing antibiotic therapy. Drug susceptibility testing was not included as a TPP requirement because it is not likely to be feasible in non-culture POC tests due to the evolving nature of typhoid resistance[8] and may make interpretation of test results too complex for users at the lowest healthcare level[29]. However, some Delphi survey respondents felt that for an RDT ever to replace blood culture it must include susceptibility testing. An RDT for diagnosis combined with epidemiological knowledge of the antibiotic sensitivity of strains, updated at intervals, could be a compromise solution.

The 2017 Global Burden of Disease study estimated that *S*. Paratyphi affected 3.4 million people annually, with 19.1 thousand deaths, compared to 10.9 million cases and 116.8 thousand deaths for *S*. Typhi.[2,3] The higher morbidity and mortality of *S*. Typhi makes it a greater public health concern, but the increasing prevalence of *S*. Paratyphi in certain regions makes it prudent for a next-generation test to detect *S*. Paratyphi as well as *S*. Typhi.[5,30,45] Delphi survey feedback noted that not being able to detect *S*. Paratyphi could undermine clinician confidence in a next-generation typhoid POC test as *S*. Typhi and *S*. Paratyphi may cause indistinguishable clinical syndromes.[4] As drug-resistant typhoid continues to spread, it may become necessary to differentiate between these two serovars prior to starting therapy due to different antibiotic susceptibility profiles.[46]

The minimum diagnostic requirement for a typhoid RDT in this TPP was ≥90% sensitivity and ≥95% specificity, with an optimum threshold of ≥95% sensitivity and ≥98% specificity. Consensus agreement was achieved in the Delphi survey for specificity but not sensitivity, reflecting the substantial variability seen in the published literature with proposed targets ranging from 80-90% for sensitivity and 90-98% for specificity.[21,36,37] The poor sensitivity of blood culture as a reference standard for typhoid diagnosis makes it difficult to accurately assess the performance of novel diagnostic tests.[47] A composite reference standard that combines multiple tests with high specificity but suboptimal sensitivity has been proposed as a possible way to improve diagnostic accuracy.[7,47] Various test combinations have been used as a composite reference standard for typhoid,[47] but respondents in this Delphi exercise advised that the adoption of a standardized composite is required before it can be included in a TPP. A standardized composite might include tests (bone marrow culture, PCR, transcriptomics etc.) that contribute to a reference standard but are not suitable for use in regular practice.

Typhoid fever is transmitted by the faeco-oral route in water and food contaminated by *S*. Typhi in human faeces and so is endemic in low-resource environments that lack access to clean water and adequate sanitation. While improvements in the infrastructure for water, sanitation and hygiene could reduce or eliminate typhoid, these are costly long-term endeavors. The newly approved Typbar-TCV vaccine may help to reduce the global burden of enteric fever caused by *S. Typhi*,[48] but an improved diagnostic test is required to accurately estimate disease incidence and facilitate targeted vaccine deployment. To have a meaningful impact on the overuse of antibiotics that has contributed to the emergence of resistance in *S*. Typhi and other bacteria, an improved typhoid POC test needs to be used as part of a treatment algorithm in conjunction with diagnostics for malaria and other causes of acute febrile illness. The isolated use of a disease-specific diagnostic test for a febrile patient may help focus treatment if positive, but a negative test may result in alternative empiric antibiotic therapy, as seen for malaria.[24,49] Drug susceptibility testing for *S*. Typhi, performed at reference laboratories, could inform local treatment algorithms based on regional antibiotic susceptibilities.[50]

This work provides the first comprehensive TPP for a next-generation POC test for typhoid fever. The main limitations of this study were the lack of consensus agreement for all TPP characteristics, and the relatively low response rate in the second round of the Delphi survey. The length of the survey may have been a barrier to completion due to the amount of time required to provide feedback on all 35 TPP characteristics. Further discussion among the typhoid community is needed to settle on the optimal target level of the health care system and the required diagnostic sensitivity. While this TPP is a first step toward improved awareness of the typhoid diagnostic needs, it is crucial to keep the conversation going and to engage global health funders, diagnostics developers, and national policy makers in the discussion on how improved diagnostic tools and related innovations can be used to improve surveillance data as well as support patient management decisions in the context of universal health care.

## Data Availability

All data can be made available.

## ACKNOWLEDGEMENTS

We thank all the participants of the 2 rounds of Delphi surveys for their invaluable contribution to shape the TPP and highlight open questions. We would further like to thank the UK aid from the British people for supporting this work.

